# Persistence of neuropsychiatric symptoms associated with SARS-CoV-2 positivity among a cohort of children and adolescents

**DOI:** 10.1101/2021.09.28.21264259

**Authors:** Victor M. Castro, Faith M. Gunning, Roy H. Perlis

**Author notes:** **Correspondence:** Roy H. Perlis, Department of Psychiatry, Massachusetts General Hospital, Simches Research Building, 185 Cambridge St, Boston, MA 02114.

## Abstract

**Background:** Post-acute sequelae of COVID-19 are common among adults. The prevalence of such syndromes among community samples of children and adolescents remains less well characterized.

**Method:** We identified all individuals age 5-18 across 2 New England health systems who had a positive SARS-CoV-2 PCR test between 3/12/2020 and 4/18/2021 and at least 90 days of follow-up visits documented in electronic health records. We identified neuropsychiatric symptoms in intervals prior to, and following, this testing using a previously-derived set of ICD-10 codes and natural language processing terms. Primary analysis examined sociodemographic features associated with presence of at least one incident (i.e., new-onset) neuropsychiatric symptom between 90 and 150 days after an initial positive test for COVID-19.

**Results:** Among 5058 children (50% female, 2.9% Asian, 6.3% Black, and 63% White; 30% Hispanic; mean age was 12.4 (IQR 8.9-15.6), 366 (7.2%) exhibited at least one new-onset neuropsychiatric symptom between 90 and 150 days following initial SARS-CoV-2 test positivity. The most common incident symptoms at 90-150 days were headache (2.4%), mood and anxiety symptoms (2.4%), cognitive symptoms (2.3%), and fatigue (1.1%). In regression models, older children, girls, those with Hispanic ethnicity, those with public versus private insurance, and those with greater overall burden of medical comorbidity were more likely to exhibit subsequent symptoms.

**Conclusion:** The prevalence of neuropsychiatric symptoms between 3- and 5-months following SARS-CoV-2 test positivity is similar to that observed in the period prior to infection. Prospective controlled studies will be needed to further refine these estimates.

## Introduction

A subset of adults with acute COVID-19 will experience persistence of symptoms beyond 3 months, a phenomenon characterized as post-acute sequelae of COVID-19 (PASC). Studies using a variety of designs, from self-report to prospective cohorts to analysis of electronic health records, have provided estimates of prevalence of such symptoms, and putative predictors of risk[1].

Neuropsychiatric symptoms are among the mostly common-observed features of PASC. Initial studies using mobile application survey-based self-report found elevated rates of neurologic symptoms beyond the first month following acute illness[2]. Two claims-based studies in the US found elevated rates of anxiety as well as memory disturbance[3][4]; in the latter study, around 1/3 of individuals received a new neuropsychiatric diagnostic code. A study in the Veterans Administration of adults diagnosed but not hospitalized for COVID-19 likewise found marked elevation in neuropsychiatric diagnoses [5]. On the other hand, a recent study that incorporated both health claims and narrative clinical notes did not identify similar risk[6], with high rates of neuropsychiatric symptoms following hospitalization *regardless* of COVID-19 status.

In school-age children, neuropsychiatric symptoms of PASC have been less well characterized. The bulk of prevalence data comes from cross-sectional surveys. Such survey studies are challenging to compare, as they vary in interval from initial diagnosis, definition of infection, and means of assessment; perhaps unsurprisingly, they have yielded widely divergent estimates of symptom prevalence, ranging from less than 5%[7] to more than 1 in 3 [8][9]. Concern about neurodevelopmental impacts have been increased by two small neuroimaging studies suggesting that, in a subset of children, objective changes indicative of neuropathology may be detectable after acute infection[10][11], although the mechanism underlying such effects is unknown.

More precise and generalizable estimates of COVID-19 sequelae among children are essential to inform decisions ranging from returning to in-person education to balancing risks and benefits of vaccination. Therefore, we used data from the electronic health records of two large health systems including academic medical centers, community hospitals, and outpatient clinical networks to generate such estimates. We applied a simple natural language processing method that we have previously shown to be more sensitive for neuropsychiatric symptoms than ICD10 diagnostic codes alone[6]. Further, we sought to identify clinical and sociodemographic predictors of symptom persistence, as these predictors may be useful in the design of prospective studies as well as targeted intervention trials.

## Methods

### Study Design and Cohort Derivation

This study applies a retrospective cohort design including individuals age 5-18 at time of first documented positive SARS-CoV-2 polymerase chain reaction (PCR) test result in the electronic health records of any of 6 Eastern Massachusetts hospitals and their outpatient affiliate networks between 3/12/2020 and 4/18/2021, and who have at least 90 days of follow-up defined by the presence of at least one subsequent note at or after 90 days or have health system PCP. The Mass General Brigham Research Patient Data Registry (RPDR) [12] was queried to generate a datamart [13] incorporating all clinical notes, ICD10 diagnostic codes, electronically-prescribed medications, and sociodemographic features collected as part of the patient registration process, including insurance type, used to capture socioeconomic status. Charlson comorbidity index[14] was calculated on the basis of all diagnostic codes present prior to positive SARS-CoV-2 test result.

The Human Research Committee of Mass General-Brigham granted a waiver of informed consent requirement for this research protocol under 45 CFR 46.116, as only secondary use of data generated by routine clinical care was required.

### Outcomes

We have previously reported an approach to capturing neuropsychiatric phenotypes that relies on a simple and transparent natural language processing (NLP) approach. In brief, a list of tokens representing individual symptoms was drawn from a symptom self-report survey [2,15] and then expanded using synonyms. The list was manually curated by the authors after reviewing randomly-selected text fragments from narrative notes. In prior work, such simple token-based concepts have been shown to correlate with neuropsychiatric measures and to predict longitudinal clinical outcomes (see, e.g., [16,17]). We excluded tokens appearing in family history and patient instruction sections of notes; otherwise, presence of at least one token in a particular domain was considered to be presence of that symptom domain. Presence of a given domain could also be identified by presence of an ICD-10 symptom code (ICD10: R*). For primary analysis, the presence of either a term, or an ICD-10 code, was considered to represent presence of a symptom domain. (See Supplemental Tables 1 and 2).

### Analysis

We defined 3 epochs preceding and following the date of the first positive SARS-CoV-2 PCR result, each spanning 60 days. Presence of a pre-existing symptom domain was defined by at least one symptom between 90 days and 15 days prior to the PCR result. (The 15-day threshold was selected to minimize the likelihood that symptoms represented acute illness). The early postacute period was defined between 30 and 90 days following PCR result, and the late postacute period was defined between 90 and 150 days following PCR result. Individuals were only considered at risk if there was at least one diagnostic code or narrative note in a given window or if the patient’s primary care physician or pediatrician is within the hospital system In addition to descriptive analysis, we utilized multiple logistic regression to characterize sociodemographic features associated with risk for presence of any of the neuropsychiatric symptoms at 90-150 days. (While superficially appealing because of the capacity to incorporate censorship at end of observed follow-up, survival analysis was not applied because, first, we sought comparable estimates of prevalence to other published work, and second, calculating time-to-event was not the aim of this investigation.)

Secondarily we examined association between concomitant medications, as markers of comorbidity, and persistence of neuropsychiatric symptoms, using crude regression models and then models adjusted for the same set of sociodemographic features. As this was an exploratory analysis, we did not correct for multiple-hypothesis testing. Medication exposure was defined using a window spanning 30 days prior to index positive PCR and 30 days following.

Analyses utilized R 4.0.3 [18].

## Results

The cohort as a whole included 5058 children age 5-18, of whom 366 (7.2%) experienced at least one neuropsychiatric symptom between 90 and 150 days following initial positive SARS-CoV-2 test. Overall, subjects were 50% female, 2.9% Asian, 6.3% Black, and 63% White; 30% Hispanic; mean age was 12.4 (IQR: 8.9-15.6). 27% had public insurance, and 95% had an identified primary care physician or pediatrician within the hospital system.

The rates of individual neuropsychiatric symptoms in time intervals before and after SARS-CoV-2 test is shown in Table 2 and Supplemental Figure 1; the most common symptoms at 90-150 days that were not present prior to infection were headache (2.4%), mood and anxiety symptoms (2.4%), cognitive symptoms (2.3%), and fatigue (1.1%). Notably, prevalence of symptoms was qualitatively similar in the equivalent 60d period prior to SARS-CoV-2 test: 9.6% had at least one symptom including 3.1%, 5.3%, 2.5%, and 1.6%, for headache, mood/anxiety, cognition, and fatigue, respectively. (For symptom co-occurrence, see Supplemental Figure 2; for overall prevalence, regardless of whether symptoms were pre-existing, see Supplemental Table 3).

**Table 1.** Features of children who tested positive for SARS-CoV-2 and did, or did not, have at least one documented neuropsychiatric symptom between 90 and 150 days later

| Characteristic | NeuroPASC-,<br>N = 4,692<sup>1</sup> | NeuroPASC+,<br>N = 366<sup>1</sup> | p-value<sup>2</sup> |
| --- | --- | --- | --- |
| **Age at index** | 12.3 (8.8, 15.5) | 13.7 (9.3, 16.5) | <0.001 |
| **Gender** |  |  | <0.001 |
| Female | 2,304 (49%) | 214 (58%) |  |
| Male | 2,388 (51%) | 152 (42%) |  |
| **Race** |  |  | <0.001 |
| Asian | 139 (3.0%) | 7 (1.9%) |  |
| Black | 297 (6.3%) | 24 (6.6%) |  |
| Other | 724 (15%) | 100 (27%) |  |
| Unknown | 561 (12%) | 38 (10%) |  |
| White | 2,971 (63%) | 197 (54%) |  |
| **Hispanic** | 1,349 (29%) | 151 (41%) | <0.001 |
| **Public insurance** | 1,211 (26%) | 133 (36%) | <0.001 |
| **Health system PCP** | 4,492 (96%) | 323 (88%) | <0.001 |
| **Charlson Comorbidity Index** |  |  | <0.001 |
| 0 | 3,999 (85%) | 293 (80%) |  |
| 1 | 674 (14%) | 65 (18%) |  |
| 2+ | 19 (0.4%) | 8 (2.2%) |  |

<sup>1</sup>Median (IQR); n (%)
<sup>2</sup>Wilcoxon rank sum test; Pearson's Chi-squared test; Fisher's exact test

**Table 2.**
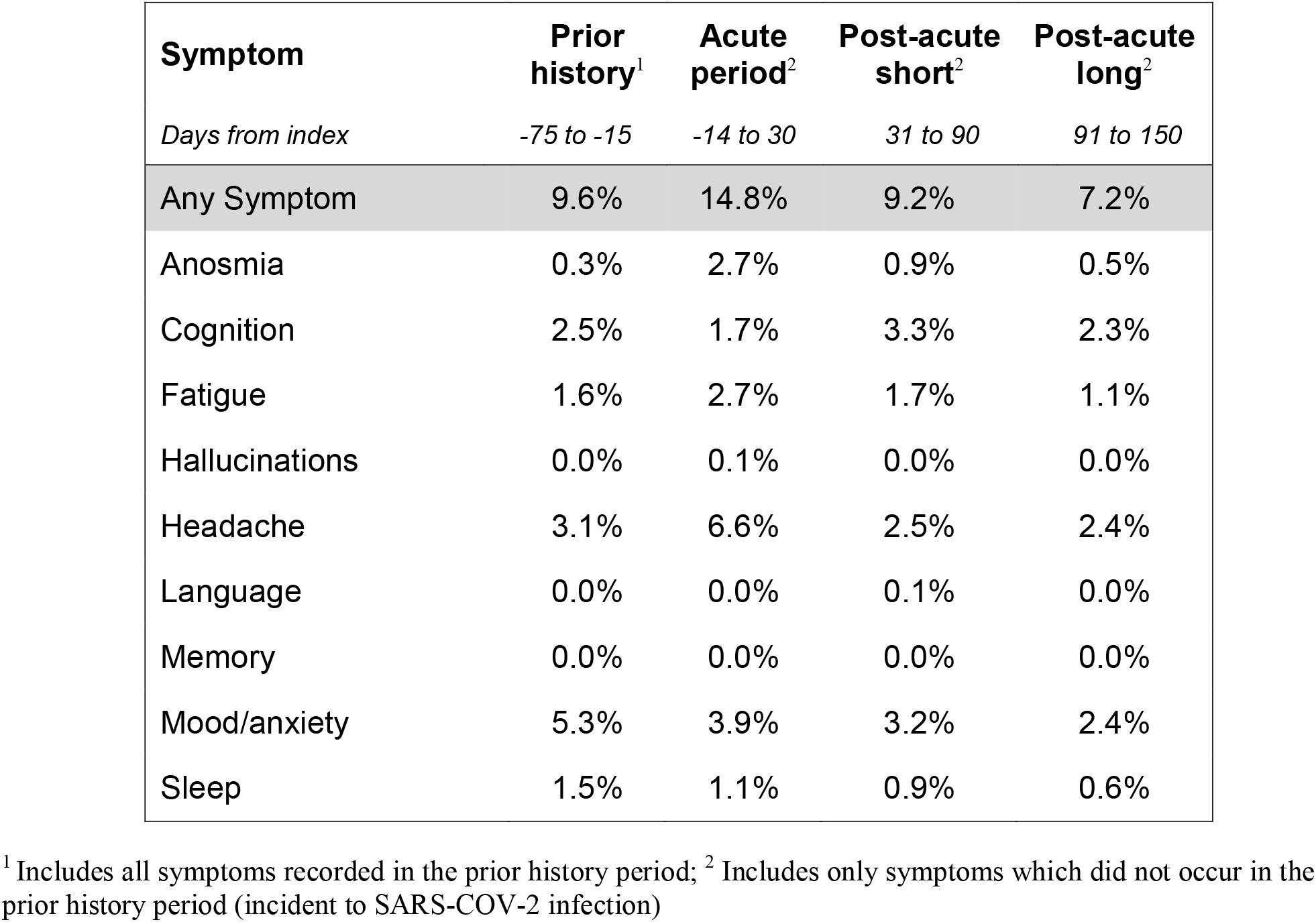
Incidence of individual neuropsychiatric symptoms in each of 4 epochs: prior to SARS-CoV-2, during acute illness, between 30 and 90 days following, and 90 to 150 days following.

In a logistic regression model (Figure 1), features associated with greater probability of symptoms at 90-150 days included older age, female gender, Hispanic ethnicity, and public insurance and Charlson comorbidity index score of 2 or greater.

**Figure 1.**
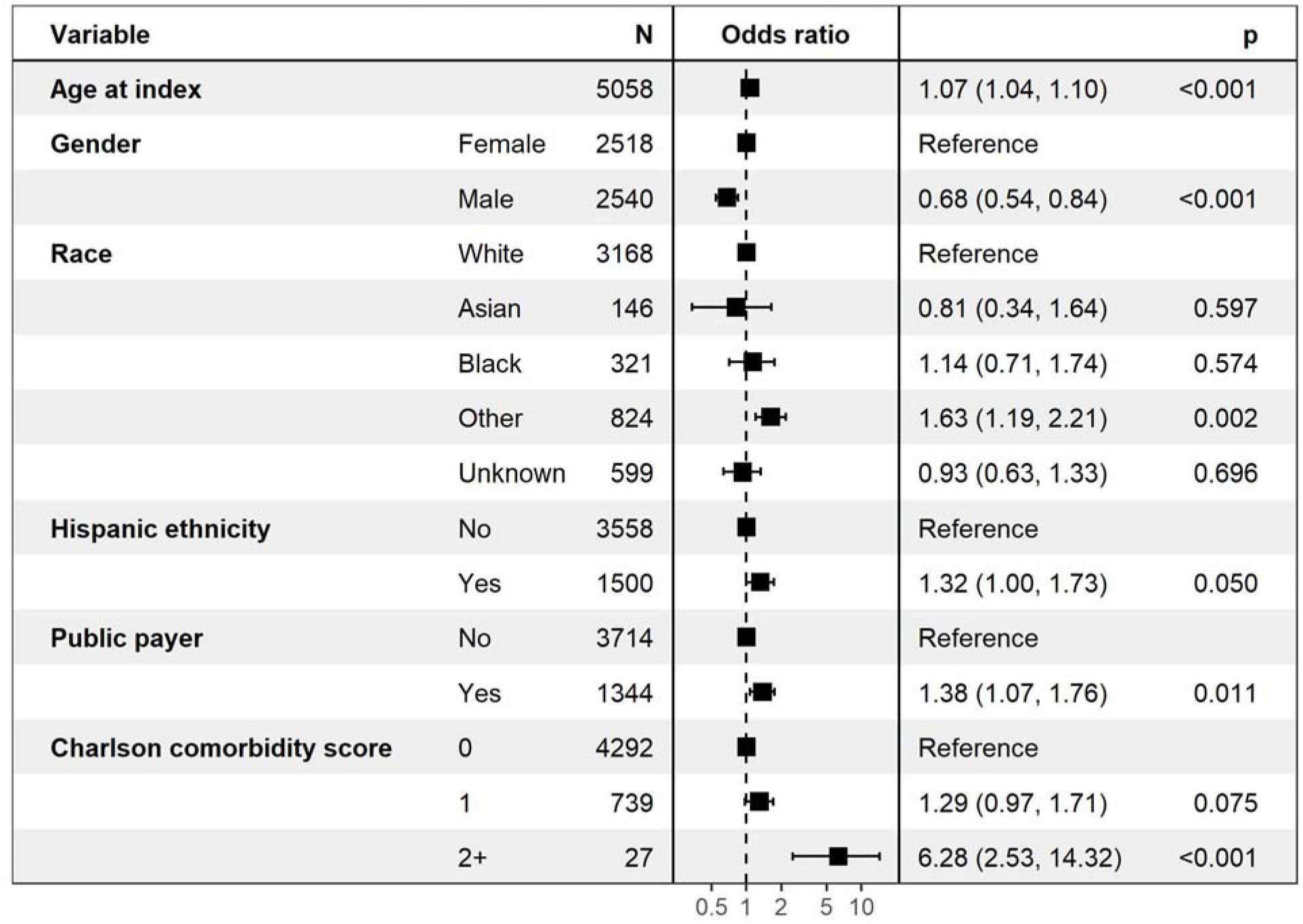
Multiple logistic regression model for presence of at least one documented neuropsychiatric symptom between 90 and 150 days after initial positive SARS-CoV-2 test

We next examined medications prescribed prior to or shortly after initial SARS-CoV-2 test, as a proxy for potential risk-increasing (or decreasing) comorbidities (Figure 2). With 224 observed medications, the nominal threshold for Bonferroni-corrected significance would be p<2.2e-4. Among those medications most associated with symptom persistence exceeding this significance threshold were metronidazole, dexamethasone, guanfacine, diphenhydramine, and clonidine (Table 3).

**Figure 2.**
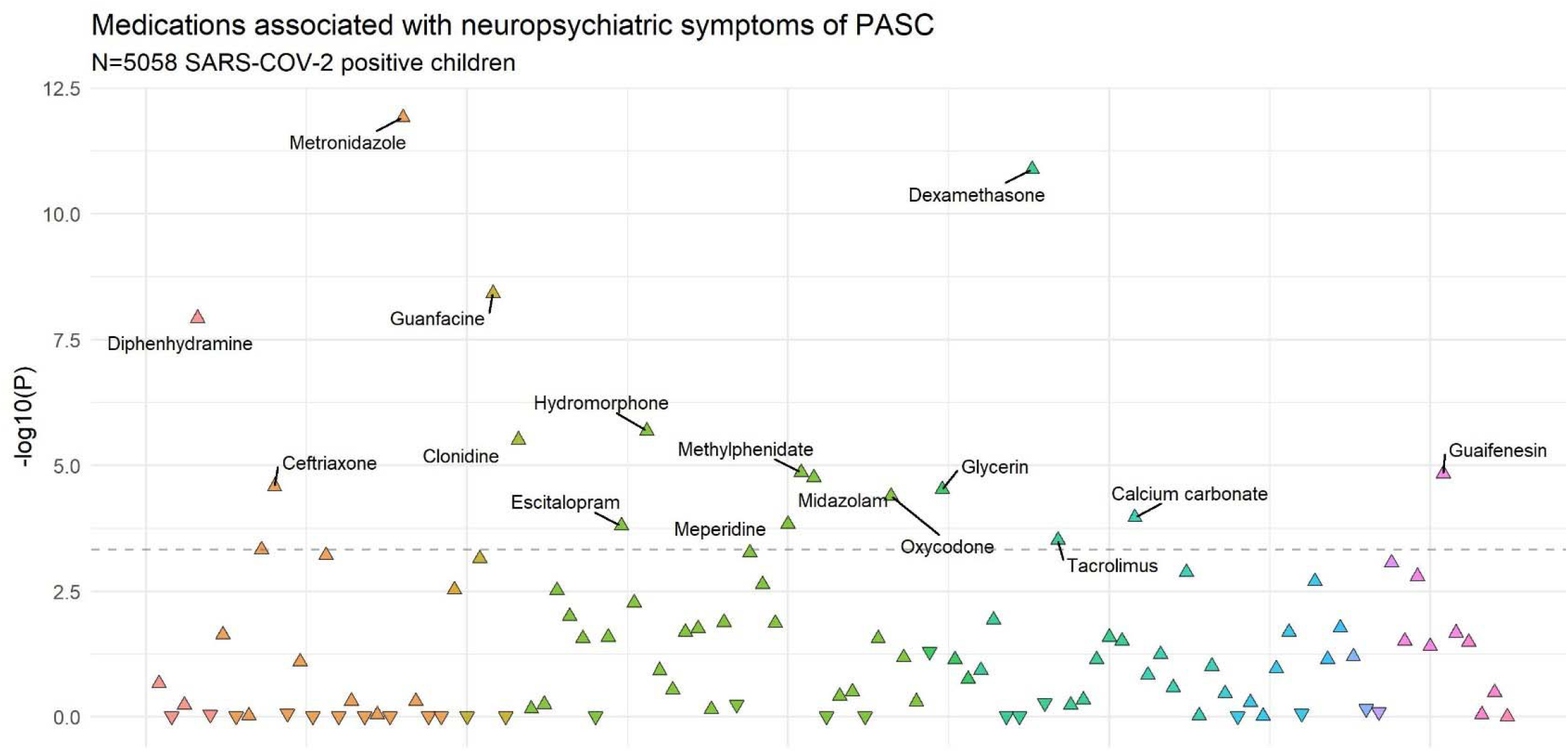
Manhattan plot of medications associated with differential likelihood of incident neuropsychiatric symptoms, based on fully adjusted multiple regression models, at 90-150 days after initial positive SARS-CoV-2 test. Dotted line indicates threshold for Bonferroni-corrected statistical significance.

**Table 3.** Medications associated with differential likelihood of neuropsychiatric symptoms between 90 and 150 days following initial positive SARS-CoV-2 test

| RxNorm ID | Medication Ingredient | OR | OR<br>95% CI<br>Low | OR<br>95% CI<br>High | p | Adj OR | Adj OR<br>95% CI<br>Low | Adj OR<br>95% CI<br>High | Adj p |
| --- | --- | --- | --- | --- | --- | --- | --- | --- | --- |
| RXNORM:6922 | Metronidazole | 12.92 | 6.66 | 25.04 | 4e-14 | 11.72 | 5.94 | 23.12 | 1e-12 |
| RXNORM:3264 | Dexamethasone | 3.45 | 2.61 | 4.58 | <2e-16 | 2.81 | 2.08 | 3.78 | 1e-11 |
| RXNORM:40114 | Guanfacine | 4.36 | 2.53 | 7.51 | 1e-07 | 5.44 | 3.10 | 9.55 | 4e-09 |
| RXNORM:3498 | Diphenhydramine | 5.45 | 3.35 | 8.88 | 1e-11 | 4.41 | 2.65 | 7.34 | 1e-08 |
| RXNORM:3423 | Hydromorphone | 10.82 | 4.64 | 25.21 | 3e-08 | 8.67 | 3.55 | 21.15 | 2e-06 |
| RXNORM:2599 | Clonidine | 3.89 | 2.27 | 6.66 | 7e-07 | 3.74 | 2.15 | 6.52 | 3e-06 |
| RXNORM:6901 | Methylphenidate | 2.43 | 1.58 | 3.75 | 6e-05 | 2.70 | 1.72 | 4.22 | 1e-05 |
| RXNORM:5032 | Guaifenesin | 19.38 | 5.44 | 68.99 | 5e-06 | 17.79 | 4.84 | 65.42 | 1e-05 |
| RXNORM:6960 | Midazolam | 5.89 | 2.77 | 12.54 | 4e-06 | 5.41 | 2.50 | 11.70 | 2e-05 |
| RXNORM:2193 | Ceftriaxone | 12.92 | 4.15 | 40.25 | 1e-05 | 11.82 | 3.74 | 37.39 | 3e-05 |
| RXNORM:4910 | Glycerin | 25.77 | 7.72 | 86.00 | 1e-07 | 14.64 | 4.15 | 51.65 | 3e-05 |
| RXNORM:7804 | Oxycodone | 5.40 | 2.56 | 11.38 | 9e-06 | 4.89 | 2.29 | 10.45 | 4e-05 |
| RXNORM:1897 | Calcium carbonate | 6.44 | 2.74 | 15.15 | 2e-05 | 5.79 | 2.38 | 14.09 | 1e-04 |
| RXNORM:6754 | Meperidine | 19.38 | 5.44 | 68.99 | 5e-06 | 13.94 | 3.57 | 54.40 | 1e-04 |
| RXNORM:321988 | Escitalopram | 4.05 | 1.97 | 8.30 | 1e-04 | 4.11 | 1.97 | 8.57 | 2e-04 |
| RXNORM:42316 | Tacrolimus | 8.59 | 3.04 | 24.26 | 5e-05 | 6.89 | 2.42 | 19.66 | 3e-04 |

## Discussion

Among more than 5,000 school-age children across 2 academic health systems, we found new-onset neuropsychiatric symptoms to be uncommon at 90-150 days following initial positive SARS-CoV-2 test, with at least one symptom observed in 7.2%. By comparison, in an equivalent time interval *before* onset of COVID symptoms, at least one symptom was observed in 9.6% of children.

Our results complement and extend prior work by capturing a large, generalizable community cohort, using clinical assessment rather than survey or other self-report. Adult investigations, summarized in a recent meta-analysis of 51 studies [1], have tended to find neuropsychiatric symptoms to be far more prevalent. However, such studies have generally focused on hospitalized populations. For example, in the same health systems from which we drew our cohorts, rates among previously-hospitalized adults were an order of magnitude greater, with fatigue in 10.9%, mood and anxiety symptoms in 8.2%, sleep disruption in 6.8%, and impaired cognition in 5.8%.

Our results are difficult to compare directly to prior studies, which have adopted divergent designs. In one of the first such reports, among 129 children assessed on average 5 months after acute infection, more than half had some PASC symptoms; common neuropsychiatric symptoms included insomnia in 18.6%, fatigue in 10.8%, and impaired concentration in 10.1%. [19]. These early findings raised substantial concern, particularly in concert with neuroimaging studies using [^18^F]-FDG-PET[10] or MRI[11] that demonstrated persistent brain changes in small subsets of children following acute infection.

Published cohort studies in children also suggest high rates of persistence. Among 518 children hospitalized for COVID-19, ∼1/4 reported persistence of symptoms at telephone follow-up on average 8 months after initial illness. In this group, fatigue was reported by 10.7% and disturbance in sleep in 6.9%[20].

This estimate reflects hospitalized children, raising the question of whether the risk may be less among community samples who on average have far less severe illness. Indeed, estimates drawn from survey data have generally been more reassuring[21]. For example, a survey-based study of ∼1700 school-age children in the UK found persistent symptoms in 4.4% beyond 28 days, and 1.8% beyond 56 days; among the most common symptoms were fatigue, headache, and anosmia, each reported in ∼4 of 5 children who were symptomatic beyond 28 days[7].

Interpreting these self-report results may be challenging, however, in the absence of a control group. The difficulty is underscored by the results of a survey of 1500 school-age children in Germany, which found individual neuropsychiatric symptoms in ∼1/3 of respondents, including impaired concentration, memory loss, mood symptoms, insomnia and headache. Notably, these rates did not differ significantly in the 12% of those who were seropositive for SARS-CoV-2 antibodies[8]. A UK cohort study relying on survey data similarly found no difference in overall mental health symptoms or functioning at 3 months following enrollment between tested children 11-17 who were seropositive, or seronegative, for SARS-CoV-2[9,22] – although that study did find seropositive participants were more likely to report 3 or more symptoms.

Taken together, these two results indicate the potential lack of specificity for COVID-19 sequelae in self-report studies, compared to clinical assessment. Therefore, we elected to examine both coded and uncoded definitions of documented neuropsychiatric symptoms. In prior work, we found that neither definition individually is sufficiently sensitive[6]. The broader capture afforded by natural language processing may avoid biases introduced by the fact that children with prior COVID-19 may receive closer follow-up than would be typical, and thus more likely to receive additional billing code diagnoses. On the other hand, capture of outcomes is less reliable than that afforded by prospective cohort studies, even though both designs pose limitations as participants are not uniformly observed and data are not missing at random.

We present analyses of concomitant medications as a means of understanding potential risk factors for persistence of neuropsychiatric symptoms, but such results must be interpreted with caution. Some of these likely represent markers of preexisting neuropsychiatric symptoms – for example, guanfacine and clonidine are often used in children for treatment of attention deficit-hyperactivity disorder, and escitalopram for depression or anxiety. Others may indicate more severe acute illness, including dexamethasone, or treatment of comorbidity during COVID-19 illness, such as metronidazole. More broadly, all such data underscore that electronic health records reflect patterns of clinical service, not regular systematic assessment – so, for example, individuals with closer follow-up for other reasons may be more likely to have neuropsychiatric symptoms documented. (For further consideration of the impact of missing data in electronic health records, see Haneuse[23]). While not a focus of this effort, the likelihood of residual confounding highlights the limitations of efforts to identify medications with off-target effects for repositioning.

Despite these challenges, more precise estimates of prevalence will be important in facilitating conversations about risks and benefits of vaccination, as well as in-person education. True estimates of the risk of infection requires consideration not just of acute impact, but of PASC risk. Even a rare outcome can have an outsize impact when it occurs early in life; studies of children with PASC symptoms up to 7 months following acute infection suggest substantial disability[24]. As such, while the low rates we observe are reassuring, further studies will be required to better understand neuropsychiatric PASC in children and adolescents.

## Data Availability

Per IRB approval, data cannot be released to outside investigators without permission.

## Author Contributions

RHP - conceived analysis, drafted and revised manuscript

FMG – interpreted results, drafted and revised manuscript

VMC - generated data set and analyzed data, revised manuscript

