## Supplemental Materials for "Persistence of neuropsychiatric symptoms associated with SARS-CoV-2 positivity among a cohort of children and adolescents"

**Table S1.** Symptom NLP Search Term

| **Symptom** | **Search term** |
| --- | --- |
| Memory | "Memory impairment" |
| Memory | "Memory loss" |
| Memory | "Getting lost" |
| Memory | "Difficulty remembering" |
| Memory | "Decreased memory" |
| Memory | "Loss of memory" |
| Cognition | "Poor Cognition" |
| Cognition | "Impaired cognition" |
| Cognition | "Cognitive impairment" |
| Cognition | "Brain fog" |
| Cognition | "Impaired Concentration" |
| Cognition | "Trouble with concentration" |
| Cognition | "Trouble Concentrating" |
| Cognition | "Difficulty concentrating" |
| Cognition | "Loss concentration" |
| Cognition | "Difficulty focusing" |
| Cognition | "Trouble focusing" |
| Cognition | "Loss focus" |
| Cognition | "Executive Functioning" |
| Cognition | "Difficulty planning" |
| Cognition | "Difficulty organizing" |
| Cognition | "Trouble planning" |
| Cognition | "Trouble organizing" |
| Cognition | "trouble Decision-making" |
| Cognition | "difficult Decision making" |
| Cognition | "difficulty making decisions" |
| Cognition | Confusion |
| Cognition | Confused |
| Cognition | "Slowed thoughts" |
| Cognition | "Slowed thinking" |
| Language | "Trouble speaking" |
| Language | "Trouble finding words" |
| Language | "Trouble word finding" |
| Language | "Difficulty speaking" |
| Language | "Difficulty finding words" |
| Language | "Difficulty word finding" |
| Language | "Can’t find words" |
| Language | "Can’t remember names" |
| Language | "Difficulty names" |
| Language | Agnosia |
| Language | "Poor word retrieval" |
| Language | "Difficulty word retrieval" |
| Language | "Difficulty writing" |
| Language | "Trouble Writing" |
| Language | "Trouble Speaking" |
| Language | "Trouble Communicating" |
| Sleep | "Broken sleep" |
| Sleep | "Restless sleep" |
| Sleep | "Decreased sleep" |
| Sleep | "Increased sleep" |
| Sleep | "Poor sleep" |
| Sleep | "Disrupted sleep" |
| Sleep | Insomnia |
| Sleep | "Restless legs" |
| Sleep | "Sleep apnea" |
| Sleep | "Vivid dreams" |
| Sleep | Nightmares |
| Headache | Headache |
| Headache | Migraine |
| Mood/anxiety | Anxiety |
| Mood/anxiety | Anxious |
| Mood/anxiety | Irritable |
| Mood/anxiety | Irritability |
| Mood/anxiety | Anger |
| Mood/anxiety | Angry |
| Mood/anxiety | Depressed |
| Mood/anxiety | Depression |
| Mood/anxiety | Apathy |
| Mood/anxiety | "Mood swings" |
| Mood/anxiety | Moody |
| Mood/anxiety | "Mood lability" |
| Mood/anxiety | Suicidal |
| Mood/anxiety | Suicidality |
| Mood/anxiety | Suicide |
| Mood/anxiety | Mania |
| Mood/anxiety | Manic |
| Mood/anxiety | Hypomania |
| Mood/anxiety | Hypomanic |
| Anosmia | "Loss Smell" |
| Anosmia | "Loss Taste" |
| Anosmia | "Change smell" |
| Anosmia | "Change taste" |
| Anosmia | "Loss olfaction" |
| Anosmia | "Change olfaction" |
| Anosmia | Ageusia |
| Anosmia | Dysgeusia |
| Anosmia | Anosmia |
| Anosmia | Dysosmia |
| Hallucinations | "Visual hallucination" |
| Hallucinations | "Auditory hallucination" |
| Hallucinations | "Hearing things" |
| Fatigue | "Poor Energy" |
| Fatigue | "Loss of energy" |
| Fatigue | Tired |
| Fatigue | Fatigue |
| Fatigue | Malaise |
| Fatigue | Tiredness |
| Fatigue | Napping |
| Fatigue | Exhaustion |
| Fatigue | Exhausted |
| Fatigue | "Decreased activity" |
| Fatigue | "Decrease activity" |
| Fatigue | "Reduced activity" |

**Table S2**. Symptom ICD Code Definitions

| **symptom** | **ICD10CD** | **ICD10CD description** |
| --- | --- | --- |
| Anosmia | R43.0 | Anosmia |
| Anosmia | R43.1 | Parosmia |
| Anosmia | R43.2 | Parageusia |
| Anosmia | R43.8 | Other disturbances of smell and taste |
| Anosmia | R43.9 | Unspecified disturbances of smell and taste |
| Cognition | R40.4 | Transient alteration of awareness |
| Cognition | R41.0 | Disorientation, unspecified |
| Cognition | R41.82 | Altered mental status, unspecified |
| Cognition | R41.840 | Attention and concentration deficit |
| Cognition | R41.841 | Cognitive communication deficit |
| Cognition | R41.842 | Visuospatial deficit |
| Cognition | R41.843 | Psychomotor deficit |
| Cognition | R41.844 | Frontal lobe and executive function deficit |
| Cognition | R41.89 | Other symptoms and signs involving cognitive functions and awareness |
| Cognition | R41.9 | Unspecified symptoms and signs involving cognitive functions and awareness |
| Fatigue | R53.81 | Other malaise |
| Fatigue | R53.82 | Chronic fatigue, unspecified |
| Fatigue | R53.83 | Other fatigue |
| Hallucinations | R44.0 | Auditory hallucinations |
| Hallucinations | R44.1 | Visual hallucinations |
| Hallucinations | R44.2 | Other hallucinations |
| Hallucinations | R44.3 | Hallucinations, unspecified |
| Language | R47.01 | Aphasia |
| Language | R47.02 | Dysphasia |
| Language | R47.1 | Dysarthria and anarthria |
| Language | R47.81 | Slurred speech |
| Language | R47.82 | Fluency disorder in conditions classified elsewhere |
| Language | R47.89 | Other speech disturbances |
| Language | R47.9 | Unspecified speech disturbances |
| Memory | R41.1 | Anterograde amnesia |
| Memory | R41.2 | Retrograde amnesia |
| Memory | R41.3 | Other amnesia |
| Mood/anxiety | R45.1 | Restlessness and agitation |
| Mood/anxiety | R45.3 | Demoralization and apathy |
| Mood/anxiety | R45.4 | Irritability and anger |
| Mood/anxiety | R45.6 | Violent behavior |
| Mood/anxiety | R45.7 | State of emotional shock and stress, unspecified |
| Mood/anxiety | R45.81 | Low self-esteem |
| Mood/anxiety | R45.84 | Anhedonia |
| Mood/anxiety | R45.850 | Homicidal ideations |
| Mood/anxiety | R45.851 | Suicidal ideations |
| Mood/anxiety | R45.86 | Emotional lability |
| Mood/anxiety | R45.87 | Impulsiveness |
| Mood/anxiety | R45.89 | Other symptoms and signs involving emotional state |
| Sleep | R40.0 | Somnolence |

**Figure S1**. Symptom incidence in 91 to 150 post-acute period

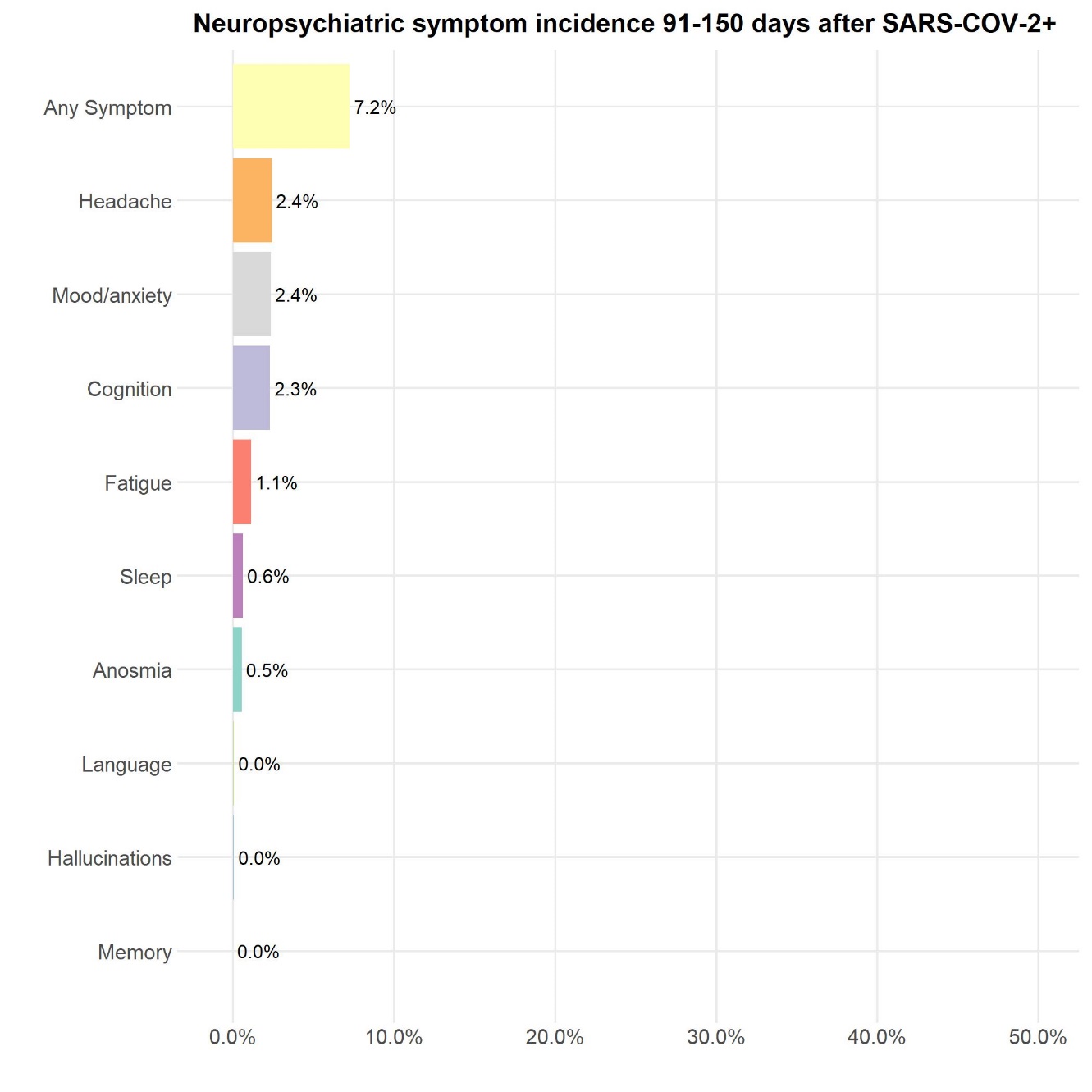

**Figure S2**. Individual frequency overlap

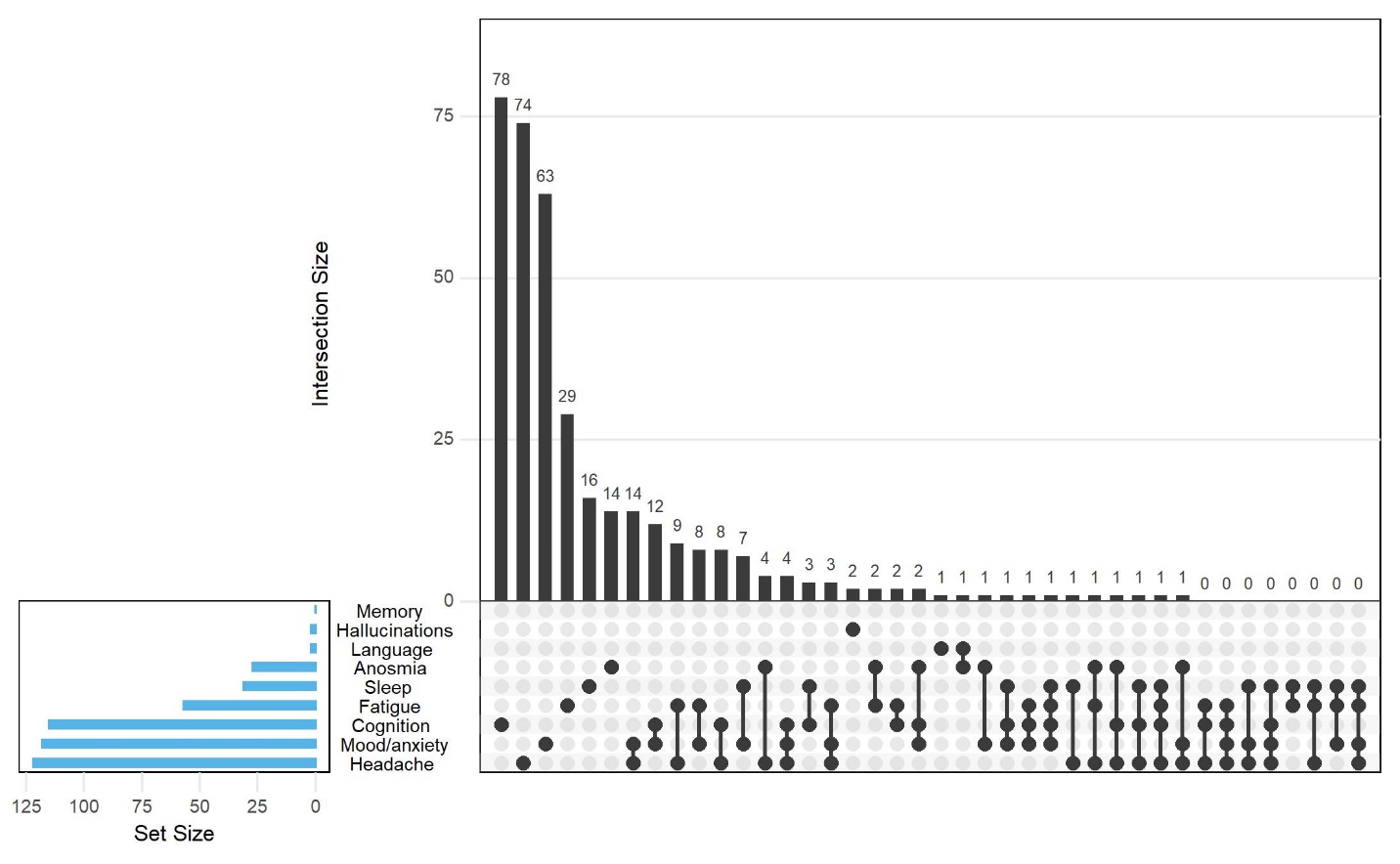

**Table S3**. Prevalence of symptoms in each period

| **Symptom** | **Prior history** | **Acute period** | **Post-acute short** | **Post-acute long** | **p-value^1^** |
| --- | --- | --- | --- | --- | --- |
| Any Symptom | 9.6% | 17.7% | 11.6% | 8.9% | <2e-16 |
| Anosmia | 0.3% | 2.8% | 1.0% | 0.6% | 0.003 |
| Cognition | 2.5% | 1.9% | 3.7% | 2.4% | 0.034 |
| Fatigue | 1.6% | 3.0% | 2.0% | 1.4% | 2e-14 |
| Hallucinations | 0.0% | 0.1% | 0.0% | 0.1% | 0.001 |
| Headache | 3.1% | 7.9% | 3.5% | 3.1% | <2e-16 |
| Language | 0.0% | 0.0% | 0.1% | 0.0% | 1.000 |
| Memory | 0.0% | 0.0% | 0.0% | 0.0% | - |
| Mood/anxiety | 5.3% | 6.5% | 5.5% | 3.8% | <2e-16 |
| Sleep | 1.5% | 1.8% | 1.4% | 1.0% | <2e-16 |

^1^ Fisher’s exact test between prior history period and post-acute long period
